# COVID-19 Serology Control Panel Using the Dried Tube Specimen Method

**DOI:** 10.1101/2021.07.07.21260101

**Authors:** William J. Windsor, Vijaya Knight, Patricia A. Merkel, Molly M. Lamb, Mario L. Santiago, Mary K. McCarthy, Thomas E. Morrison, Ross M. Kedl, Ashley Frazer-Abel, Kejun Guo, Gillian Andersen, Leah Huey, Bradley S. Barrett, Jessica M. Colón-Franco, May C. Chu

## Abstract

We used the dried tube specimen (DTS) procedure to develop the COVID-19 Serology Control Panel (CSCP). The CSCP contains five well-characterized SARS-CoV-2 pooled plasma samples made available for labs around the world to compare test kits, use for external quality assurance, harmonize laboratory testing, and train laboratory workers.

**Article Summary Line:** The dried tube specimen system is a highly effective and resilient way to provide laboratories with well-characterized serology materials. The CSCP can help clinical laboratories inform their choice of diagnostic test to supplement clinical diagnoses of SARS-CoV-2 infection.

## Text

A standardized panel composed of well-characterized plasma/serum specimens can bridge serosurveillance studies, compare test kits, serve as external quality assurance, harmonize tests that measure vaccine efficacy, be used as a training tool and be used for post-market assurance. The standardized panel can be shared by a global network to link studies and enable inclusive analysis for a variety of use cases as mentioned and, more importantly, a standardized control panel can provide long-term quality performance monitoring as reagents and production batches change.

We have established a COVID-19 Serology Control Panel (CSCP) using the dried tube specimen (DTS) protocol^1^ so that the panel can be shipped globally without a cold-chain, thus allowing equitable sharing of the materials in all resource settings. Identifying the appropriate test kit for a use-case is complicated, with >120 SARS-CoV-2 serological test kits listed by the Food and Drug Administration (FDA) under Emergency Use Authorized (EUA) and/or registered with the Conformitè Europëenne (CE)-marked European market. In addition, we can deduce from the SeroTracker^2^ list that there are at least as many research use only (RUO) tests being used in clinics and research laboratories as well. With this unprecedented number of serological testing platforms and algorithms, it is imperative that we prioritize the quality calibration of test kits and platforms. This will ensure that results are meaningful and can be compared across the hundreds of seroprevalence studies being undertaken.^3^ This is especially important for testing in low resource settings, where immunological testing is more likely to be used than other diagnostic test formats.

## The Study

Nine highly-reactive COVID-19 convalescent plasma samples collected between March-May 2020 were screened by SARS-CoV-2 spike (S) reporter viral particle neutralization assay (RVPN), with neutralization range of 1:640 to >1:10,240 (Vitalant Research Institute, VRI; San Francisco, CA) and by Ortho VITROS Anti-SARS-CoV-2 Total immunoglobulin assay (Ortho-Clinical Diagnostics, Inc. Rochester, NY) against the S subunit (S1) protein (Table 1).^4^ These nine COVID-19 convalescent plasma and one pre-2019 human plasma were sourced from VRI, Denver, CO and certified to be blood-borne pathogen-free. This protocol has a University of Colorado-Denver (CU) Human Subjects Research Waiver (Protocol 20-0711).

**Table 1:**
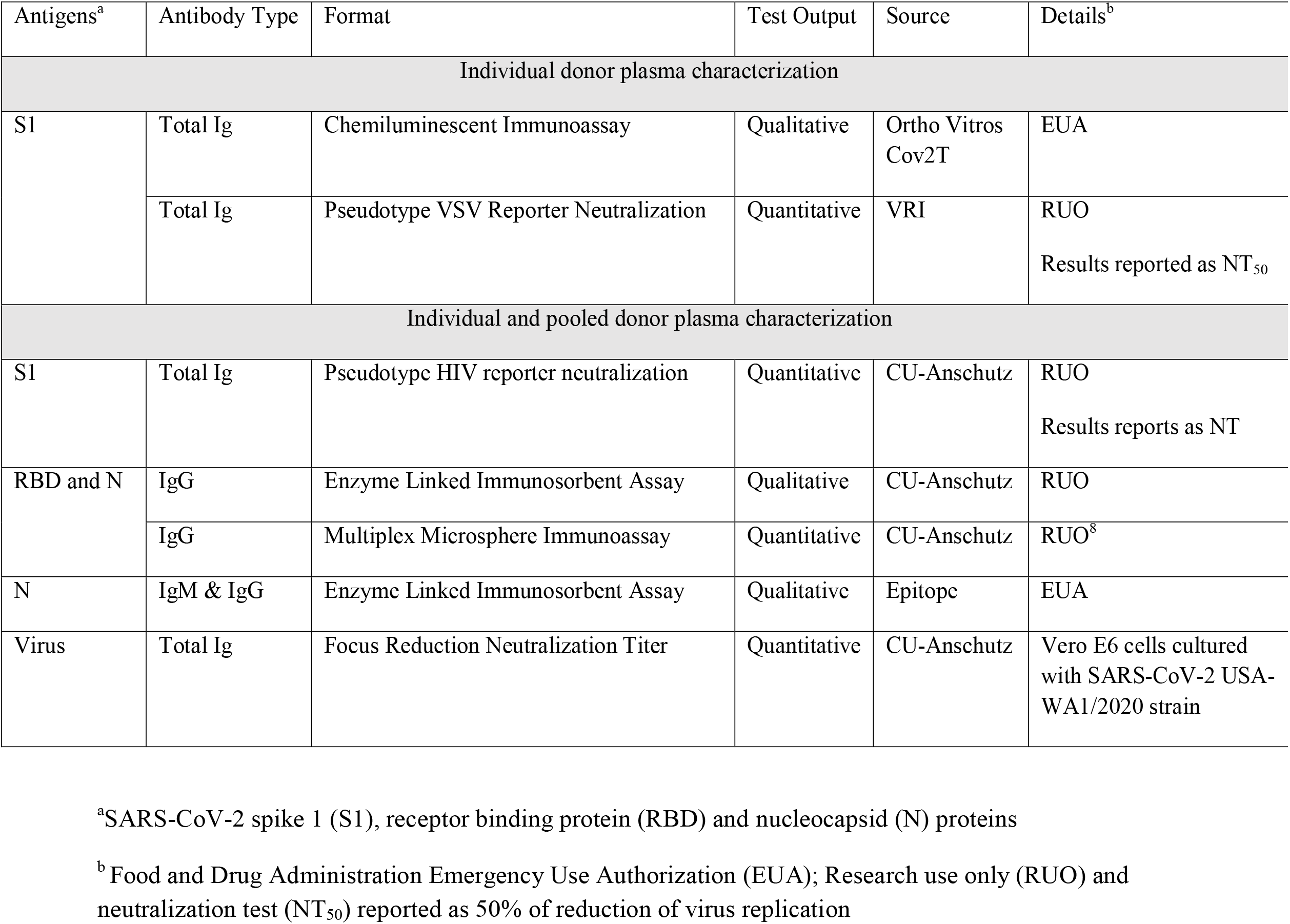
Test platforms used to characterize the CSCP samples

| Antigens<sup>a</sup> | Antibody Type | Format | Test Output | Source | Details<sup>b</sup> |
| --- | --- | --- | --- | --- | --- |
| Individual donor plasma characterization |  |  |  |  |  |
| S1 | Total Ig | Chemiluminescent Immunoassay | Qualitative | Ortho Vitros Cov2T | EUA |
|  | Total Ig | Pseudotype VSV Reporter Neutralization | Quantitative | VRI | RUO<br>Results reported as NT<sub>50</sub> |
| Individual and pooled donor plasma characterization |  |  |  |  |  |
| S1 | Total Ig | Pseudotype HIV reporter neutralization | Quantitative | CU-Anschutz | RUO<br>Results reports as NT |
| RBD and N | IgG | Enzyme Linked Immunosorbent Assay | Qualitative | CU-Anschutz | RUO |
|  | IgG | Multiplex Microsphere Immunoassay | Quantitative | CU-Anschutz | RUO<sup>8</sup> |
| N | IgM & IgG | Enzyme Linked Immunosorbent Assay | Qualitative | Epitope | EUA |
| Virus | Total Ig | Focus Reduction Neutralization Titer | Quantitative | CU-Anschutz | Vero E6 cells cultured with SARS-CoV-2 USA-WA1/2020 strain |

<sup>a</sup>SARS-CoV-2 spike 1 (S1), receptor binding protein (RBD) and nucleocapsid (N) proteins
<sup>b</sup> Food and Drug Administration Emergency Use Authorization (EUA); Research use only (RUO) and neutralization test (NT<sub>50</sub>) reported as 50% of reduction of virus replication

We tested the 10 donor samples at University of Colorado laboratories (CU Labs) to determine each sample’s reactivity to Spike (S), Nucleocapsid (N) and Receptor Binding Domain (RBD) with five SARS-CoV-2 serology methods (Table 1)^5-8^. We pooled three samples that represented the highest reactivity to S, N and RBD in a 1:1:1 ratio. The undiluted pool served as the high reactive (HR) sample and the pre-2019 served as the non-reactive (NR) in all the assays. The low reactive (LR) pool was prepared as a 1:4 dilution of the HR using the NR sample as the diluent. Then the three samples—HR, LR and NR—were evaluated by CU Labs to ascertain their reactivity by dilution and mixing, and evaluated again post-drying (Figure 1a-1e).

**Figure 1.**
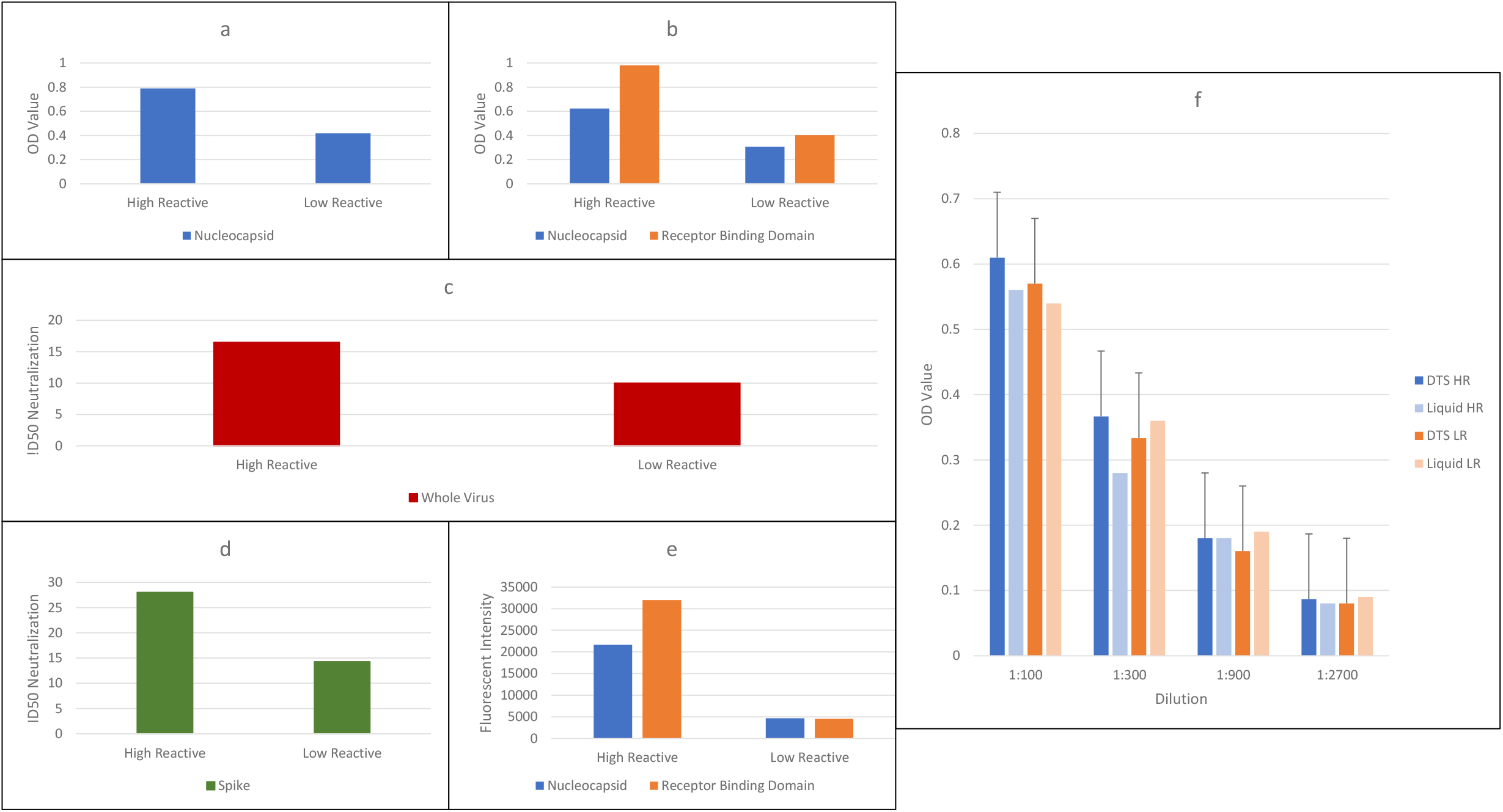
Overview of CSCP characterization results: Evaluation of pooled CSCP high reactive (HR) and low-reactive (LR) samples (a-e) and reactivity of pooled HR and LR samples before and after drying (f) Reactivity results of the high and low pooled plasma samples prior to drying. Each sub-figure represents a separate test platform and its corresponding results value are expressed on the y-axis: (a) Epitope ELISA measuring anti-Nucleocapsid; (b) CU-Anschutz developed ELISA; (c) Focus Reduction Neutralization Test with SARS-CoV-2 USA-WA1/2020 strain; (d) Pseudo-virion Neutralization Test and (e) Multiplex Microsphere Immunoassay.^8^ (f) Reactivity of pooled HR and LR samples before and after drying. Triplicate DTS and the pre-dried pool (liquid) sample were quantitatively assessed in dilution series as noted on the x-axis and OD value of the Epitope test results on the y-axis.

We followed the DTS protocol^1^ to prepare the samples, mixing in 0.1 % green food dye for better visualization. We aliquoted 20 μl in a 2 ml Sarstedt (NC9180825; Fisher Scientific, USA) and left the open tube to dry overnight under a HEPA filtered laminar flow hood. The tubes were capped and stored at 4° C during the CSCP kit assembly process and stored at -20° C afterwards to preserve the integrity of the samples for longer term storage. Each of the DTS was rehydrated by addition of 200 μl phosphate buffered saline with 0.2% Tween (PBS diluent) then tested in tandem with its corresponding pre-dried (liquid) sample to ensure that there was no change in reactivity due to the drying, assembly, and rehydration steps (Figure 1f).

We evaluated the CSCP long term temperature stability by storing kits continuously at: -20°C, 4°C, 25°C, 37°C and 45°C for 1 week, 2 weeks, 1 month, 3 months, 6 months and 1 year. The optimal stability of the CSCP for up to 6 months is between -20°C and 25°C (Figure 2) with loss of reactivity after 2 months at 37°C and is non-reactive at 45°C (data not shown). In the 6-months’ evaluation, we compared the CSCP DTS samples with the World Health Organization SARS CoV-2 Serology International Standard (WHO IS)^9,10^ and found the HR sample runs are in the same reactive range (Figure 2).

**Figure 2.**
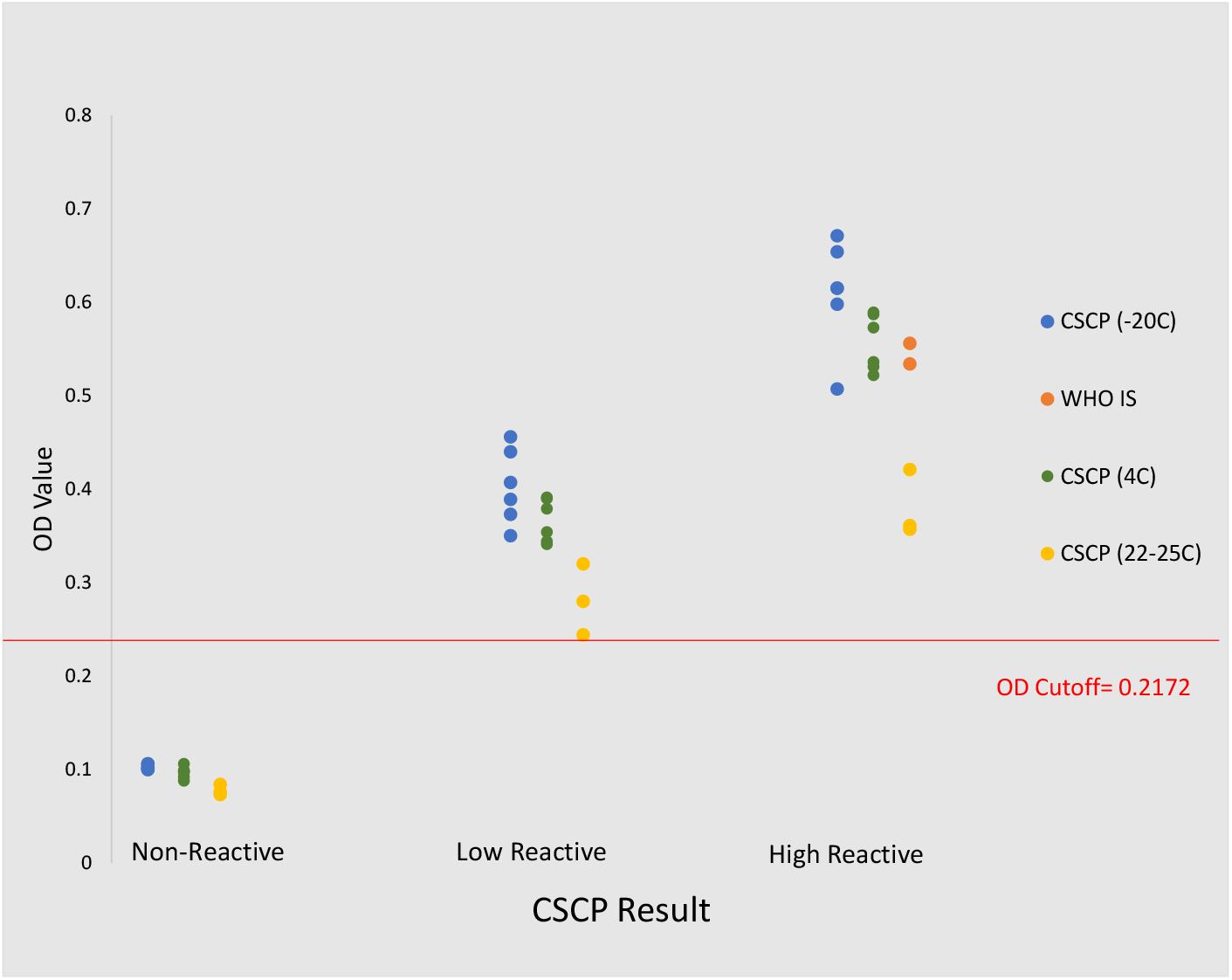
CSCP stability at 6 months, -20° C, 4° C and 22-25° C and inclusion of the WHO IS reactivity Results displayed were triplicate DTS sets incubated continuously at -20° C (blue dot), 4° C (green dot), and 22-25° C (yellow dot) for 6 months, reconstituted per protocol with PBS diluent and tested using the Epitope ELISA assay. Result values are displayed as OD measurement on the y-axis. The red line is the OD cut-off value. The WHO IS was run in duplicate (red dot) concomitantly with the triplicate DTS samples.

A CSCP kit contains five DTS samples (blinded), 200 μl of PBS diluent, a 0.5 ml calibrated disposable micropipette, a dry silica pack, a printed DTS rehydration work aide and a copy of the report form. Contents are sealed in a 1.5×9 inch mailing tube. Interested users fill out a <u>CSCP request form</u> and each request is reviewed, assessed and approved before being the requested CSCP kit is shipped at ambient temperature with no cold packs. Users then use the kit according to their serology assay requirements and report the results electronically (<u>CSCP Result Form</u>) or email a copy to. We then send the user a results form that provides a comparison and interpretation of their results against the assigned value for the DTS samples that they received.

As of June 2021, the CSCP has been shipped to multiple sites in Australia, Africa, Southeast Asia, North and South America, and Europe. Table 2 shows a summary of the 30 user results reported to us thus far, with CSCP concordance of the HR, NR, and LR being 97%, 93%, and 65% respectively. The LR samples were expected to be missed in the less sensitive assays (Table 2). Users have shared with us some logistical challenges encountered, including kits that were in transit for up to 3 months under harsh conditions (1 user) and some kits held for months before use (2 users), without change in the expected results. One user has reported issues with incomplete reconstitution and 2 users had problems uploading results to the website, all of which are under review for corrective actions.

**Table 2.** Concordance of user results with CSCP kits^a^ containing NR, HR and LR

| Test format that the CSCP user reported on: | Non-Reactive |  | High Reactive |  | Low Reactive |  |
| --- | --- | --- | --- | --- | --- | --- |
|  | N | % | N | % | N | % |
| Chemiluminescence Immunoassay | 4/4 | 100 | 2/2 | 100 | 2/4 | 50 |
| Chemiluminescent Microparticle Immunoassay | 8/8 | 100 | 4/4 | 100 | 0/8 | 0 |
| Electrochemiluminescence Immunoassay | 6/6 | 100 | 3/3 | 100 | 3/6 | 50 |
| Enzyme-Linked Immunosorbent Assay | 14/18 | 78 | 9/9 | 100 | 18/18 | 100 |
| Indirect Fluorescent Antibody Tests | 2/2 | 100 | 0/1 | 0 | 0/2 | 0 |
| Lateral Flow Assay | 18/20 | 90 | 9/10 | 90 | 14/20 | 70 |
| Multiplex Microsphere Immunoassay | 2/2 | 100 | 1/1 | 100 | 2/2 | 100 |
| Total | 54/60 | 90 | 28/30 | 93 | 39/60 | 65 |

<sup>a</sup>Each CSCP kit contains 5 coded DTS containing non-reactive, (NR), high reactive (HR) and low reactive (LR) samples with some of them repeated for in-test precision. To obtain interpretation of the coded samples, users will have to submit their results on the test platform they used, then receive a report of their correct/incorrect interpretation. The columns N are the numbers of samples that were interpreted correctly (numerator)/total of samples tested for that test platform (denominator) and the % is the number of the total samples tested that were correctly interpreted. “0” denotes none of the samples tested by users with that particular test format were concordant with the CSCP results.

## Conclusions

Quality assurance is foundational for validating methods, external quality assurance, training, and inter-/intra-laboratory comparison of serologic tests.^3,11-13^ As more of the world’s population receive COVID-19 vaccines, highly accurate and reliable SARS-COV-2 serology testing is the primary method to assess vaccine efficacy.^14^ Many commercial and laboratory developed tests react with a range of antigen targets, making it difficult to compare results in the absence of a common set of reference materials. Widespread use of the CSCP for comparison of SARS-CoV-2 tests will help laboratories interpret and gain confidence in their results while deterring laboratories from using poorly performing tests. Additionally, the CSCP will help clinical laboratories inform their choice of diagnostic test to supplement clinical diagnoses of SARS-CoV-2 infection.

With this use in mind, our next step is to harmonize CSCP and other available serology reference materials by validating them as secondary standards to the WHO IS. This would provide an inferential link to WHO IS and give broader access of validated reference materials to be used in comparing and evaluating test kits performances in use-cases already cited. The DTS system is also flexible enough to accommodate additional samples to reflect current pandemic situations, such as post-vaccination and convalescent samples from persons infected with SARS-CoV-2 variants.

## Data Availability

The Data are available

## Acknowledgments

The authors wish to acknowledge the administrative assistance of Mary Moua and Linda S. Gabel. We are grateful to Susan Fink, Tara S. Givens, Suellen Nicholson, Andy Schnaubelt for evaluating CSCP kits in their laboratories prior to kit release. This project was funded by the Bill and Melinda Gates Foundation, Investment ID INV-006307 grant.

## Disclaimers

The authors have no conflict of interest to declare.

## Author Bio

Mr. Windsor is a senior research associate with the Center for Global Health at the Colorado School of Public Health, Anschutz Medical Center, Aurora CO, USA. He is a public health projects manager including this CSCP project.

---

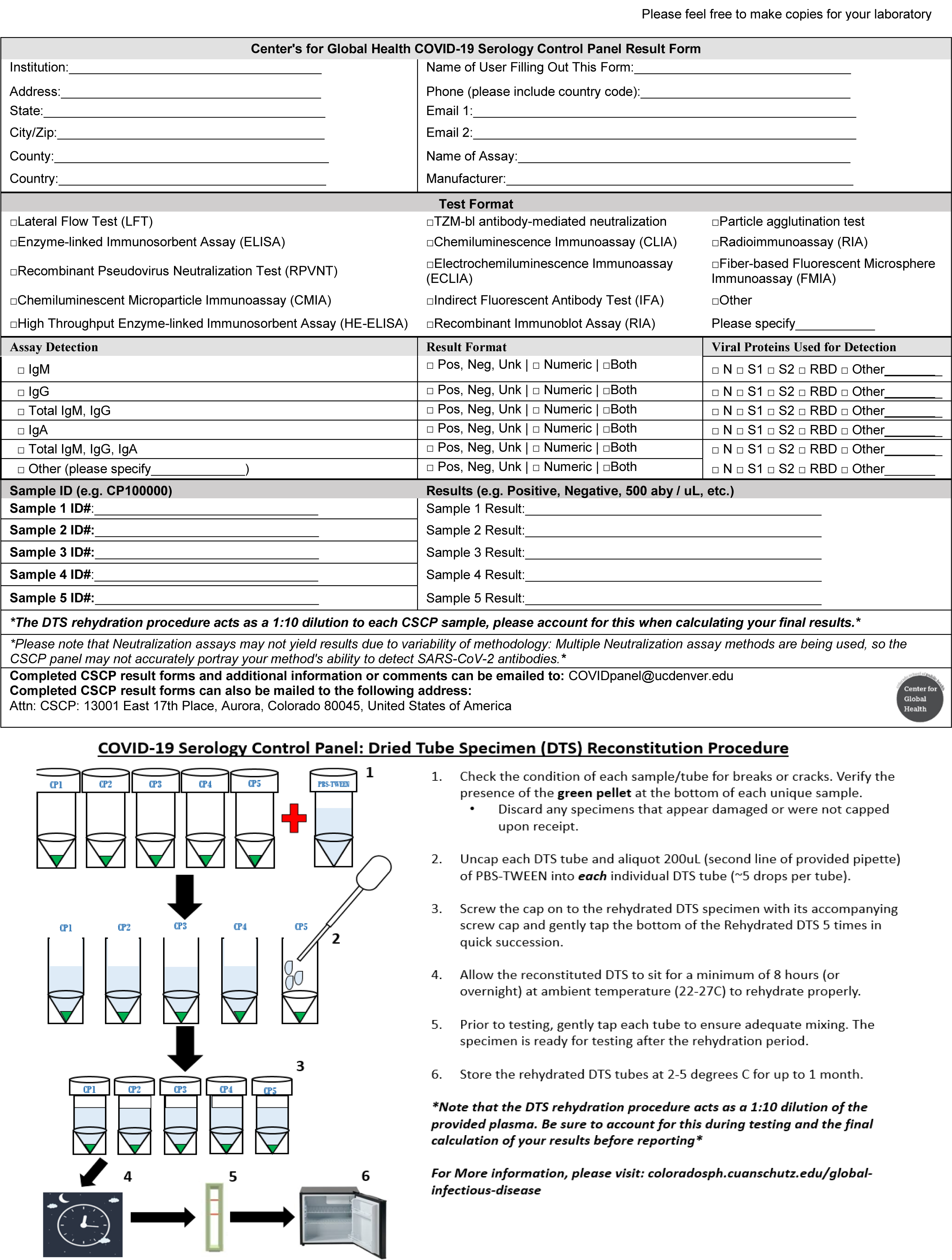

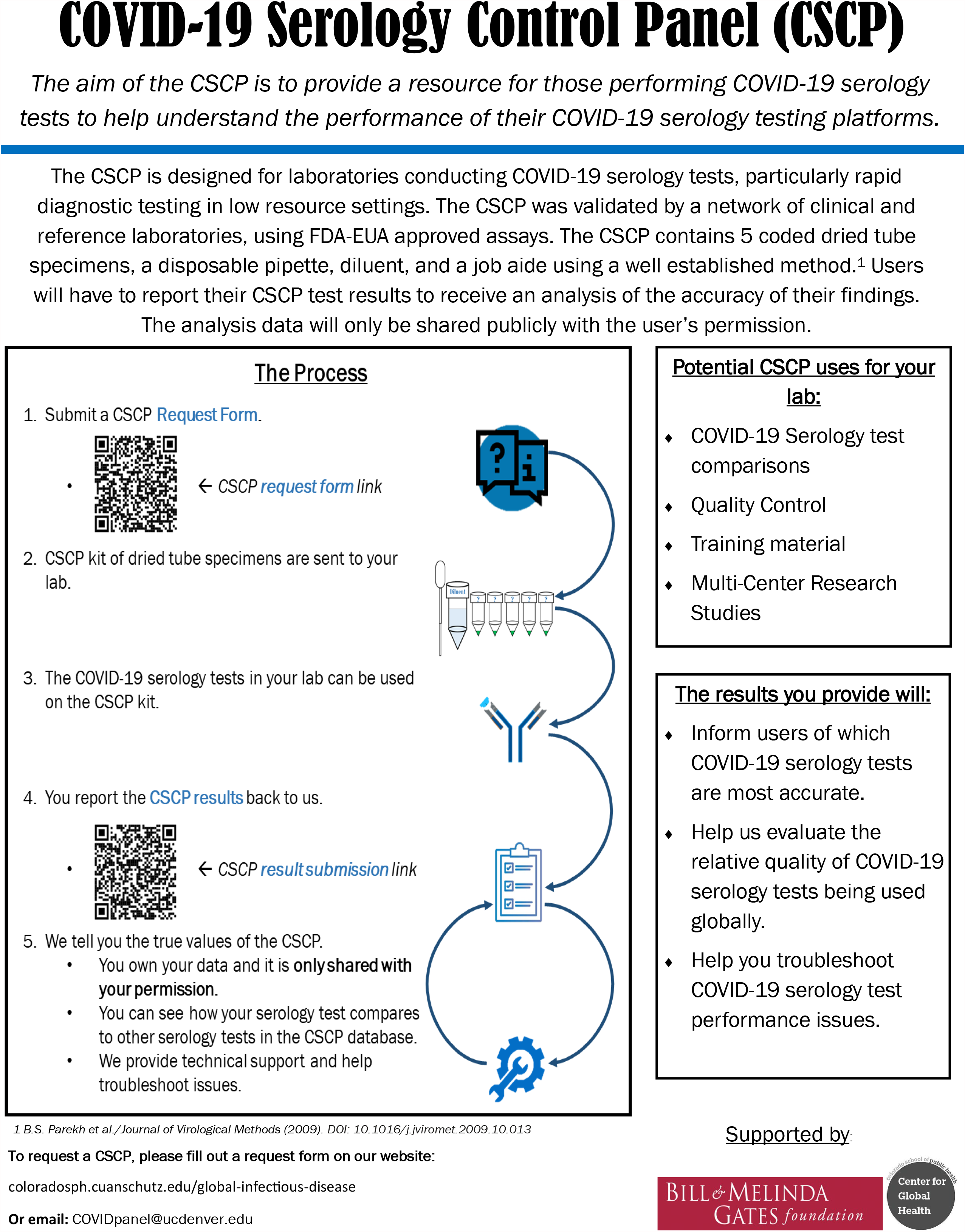

